# Assessment of Psychometric Vigilance on Neonatal Transport: A Western Australian Experience

**DOI:** 10.1101/2024.03.07.24303951

**Authors:** Alexander Wilson, Kylie McDonald, Matthew N. Cooper, Paul Stevenson, Jonathan Davis, Sanjay K. Patole

**Affiliations:** Child and Adolescent Health Service, Perth Children’s Hospital, 15 Hospital Ave, Nedlands, WA, 6009, Australia; Newborn Emergency Transport Service Western Australia, Perth Children’s Hospital, 15 Hospital Ave, Nedlands, WA, 6009, Australia; Telethon Kids Institute, 15 Hospital Ave, Nedlands, WA, 6009, Australia; University of Western Australia, 35 Stirling Highway, Crawley, WA 6009, Australia

## Abstract

**Objectives:** To assess whether undertaking retrieval was associated with fatigue independent of sleep and circadian disruption.

**Background:** Fatigue is associated impaired clinician performance and safety. The association between shift work, sleep deprivation and circadian disruption is well established. No studies have specifically assessed the independent effect of the retrieval environment on fatigue.

**Method:** Medical and nursing staff of the neonatal retrieval team were prospectively recruited over a 12-month period. Simple reaction times (RT) were recorded at the start and end of a day shift using a validated 3-min Psychometric Vigilance Test (PVT).

**Results:** End of shift RT increased by 6.38ms (95% CI: -2.17 to 14.92ms, p = 0.149) when retrieval was undertaken. A 1ms increase in RT increased the odds of being in a subjective sleepy category by 0.57% (log odds: 0.0057, 95% CI: 0.0036 to 0.0078). Consuming caffeine during the shift increased mean RT by 16.26 ms (95% CI: 4.43 to 28.1 ms, p <0.01).

**Conclusion:** The 3-min PVT was found to be an easy method of objectively assessing fatigue in the retrieval setting. The effects of caffeine consumption on RT warrants further investigation.

**Funding/Support:** This research did not receive any specific grant from funding agencies in the public, commercial, or not-for-profit sectors.

**Highlights:**

- Fatigue is associated with impaired clinician performance and safety.
- The 3-min PVT can easily be used as a proactive measure of performance in the retrieval setting, which in combination with fatigue surveys and sleep monitoring can form part of a comprehensive fatigue risk management system.
- Reaction Times were observed to be slower among participants who went on a retrieval during their shift.
- Consuming caffeine during the shift increased mean RT.

## Introduction

Fatigue during shiftwork is a concern in healthcare workers, especially in critical care and retrieval services ^1^. A high level of vigilance is essential to detect deterioration in the patient’s condition ^2^. Fatigue following sleep deprivation is associated with prolonged reaction time, decreased vigilance, perceptual and cognitive distortions, and changes in affect ^3^. There is little data on fatigue in retrieval medicine ^4-8^. Prolonged patient retrievals are associated with fatigue due to exposure to environmental factors including lower oxygen concentration inflight, reduced ambient light, vibrations, and low relative humidity ^2^. In one study fatigue accounted for 2.2% of human based contributing factors in patient transport incidents ^9^. To our knowledge there are no studies reported on assessment of fatigue in neonatal retrieval staff. Based at Perth Children’s Hospital (PCH), the Newborn Emergency Transport Service of Western Australia (NETS WA) provides the sole regional neonatal retrieval service for the state. High-risk neonates are transferred to the PCH NICU for surgical or specialist services or the King Edward Memorial Hospital for preterm care. In 2021 there were a total of 1280 neonatal retrievals in WA. Retrievals from the Perth metropolitan area were by road and transport times range from 1-4 h. WA is approximately the same size as Western Europe. Retrievals from outside the Perth metropolitan area often require retrieval by air, with duration ranging from 6-12 h. There is currently no accepted method for assessing fatigue in the retrieval setting. Psychometric Vigilance Testing (PVT) is a validated and extensively used method to objectively measure fatigue-related changes in cognitive functions in a wide variety of settings ^1,10-13^. This is assessed by measuring simple reaction times (RT) to a visual stimulus. The aim of this study was to determine whether undertaking retrieval was associated with slower RT. We hypothesized that increased exposure to the retrieval environmental would be associated with slower RT.

## Methods

### Design, setting, and participants

This prospective observational study was conducted over 12 months (March 2020 – March 2021) and involved the medical and nursing staff of NETS WA. NETS WA operates two retrieval teams which are available 24 hours a day, 7 days a week. Each team consists of one neonatal specialist trainee doctor and a specialist nurse. Participants were recruited face to face and via email for obtaining written consent before participating in the study. Exclusion criteria included any previously diagnosed sleep disorder or medical condition which may not allow a participant to wear a wrist device during the night. This research complied with the tenets of the Declaration of Helsinki and was approved by the Child and Adolescent Health Services Human Research Ethics committee (RGS0000003707) before commencing the study.

### Primary outcomes

The primary outcome was fatigue, defined and measured as reduced performance using PVT with simple RT.

#### PVT

The 3-minute PVT was measured using the Sleep-to-Peak app (https://www.neurosummum.com/en/; Neuro Summum, Quebec, Canada) on an iPad^®^ (Apple, California, USA) in the office setting. It assesses the RT in milliseconds to a visual stimulus, which is tapping the screen of the iPad after the appearance of a sun over a blue screen. All participants received training on the Sleep-2-Peak app. The Sleep-2-peak app was tested on participants 3 times as recommended ^14^. An RT of <100 ms was considered a false start and not included in the final analysis of the PVT data, except for when calculating performance score. Standard PVT outcome variables measured included: mean RT, median RT, 10% fastest, and the 10% slowest RRT, false starts (RT <100ms), number of lapses (>355ms in the 3min test), performance score, (1 minus the number of lapses and false starts divided by the number of valid stimuli including false starts) ^11,15,16^. Reduced duration, quality and quantity of sleep is one of the main contributors to fatigue and was considered to be the main confounding variable of this study ^17-20^. Accurate measurement of participant’s sleep was therefore important to include and adjust for in the analysis. Sleep actigraphy is a validated objec5 tive method for assessing sleep with high accuracy, compared with polysomnography ^21,22^. The Stanford Sleepiness Survey is a validated assessment of subjective sleepiness, with a score from 1 to 7 ^23,24^. This assessment was used to compare of whether subjective sleepiness correlates with objective PVT measurements.

### Secondary outcomes

These included: (1) Assessment of sleep duration using sleep actigraphy. This wearable wrist device contains an accelerometer that records movement. (2) Assessment of Stanford Sleepiness Survey (SSS) score^25^ (subjective sleepiness). (3) Other potential confounding variables including the duration of transport, caffeine intake, and clinical role (doctor vs. nurse).

### Data collection

Only day shifts were used to minimize the effects of circadian disruption and sleep deprivation. The NETS shifts with no retrievals undertaken were used as controls. To avoid introducing bias if one team is particularly busy or quiet, only one team member could be assessed at a time (i.e., nurse or doctor). PVT data was collected within a 2hr window at the start (baseline) and at end of the shift. Each participant was assessed over 4 shifts, considering the varying levels of activity on different shifts. Data on retrieval duration, frequency and mode of transport were recorded at the end of each shift.

Due to the 24/7 nature of retrieval shift work, participants were required to directly enter their own data via a Redcap email survey. The primary email survey included age, gender, clinical role, years of clinical experience, whether they had any previous fatigue management education, shift duration and estimated sleep (hrs) the night before each shift. New email surveys were sent out at the start and end of each shift so that participants could record their reaction times (measured via the sleep-to-peak app (Neruo Summum, Quebec, Canada), subjective sleepiness (using the subjective sleepiness scale) and whether caffeine was consumed. At the end of the shift the participants recorded the number of retrievals during their shift, duration (hours) of retrieval, mode of transport (fixed wing or road) and priority tasking geographical location (rural or metropolitan). Priority tasking is assigned based on patient acuity, with P1 being time critical, P2 Urgent, P3 Non-urgent and P4 elective. Research grade sleep actigraphs were worn the night preceding each day shift being studied using the Actiwatch Spectrum Plus (Phillips Respironics, Amsterdam, Netherlands). Sleep duration increments was measured to nearest 30 minutes. Participants were instructed to use the actigraphs to mark the time they intended to sleep and once they woke up (objective), and to record the number of hours they thought they had slept (subjective). Actigraph data was downloaded and analysed by two investigators, using Phillips Actiware software (Amsterdam Netherlands), and data from the clinician reports generated were entered by the investigators into the Redcap database.

### Statistical analysis

Sample size estimates, to inform study design, were based on data reported in Brunet et al 2017 ^14^, where a 10% increase in reaction time was observed in the sleepy state mean RT versus the alert state mean RT. We estimated that at least 11 participants were required to give >90% power to detect a minimum difference of 10% in RTs (two-tail difference between two dependent means (matched pairs); alpha 0.05). To allow for missing data owing to potential technical difficulties (and/or transfer of data issues), we aimed to recruit 20 participants. Prior to analysis, all continuous values were centered (shifted such that their mean was equal to 0) to reduce potential collinearity issues.

Linear mixed effects regression ^25^ models were used to quantify the association between the independent variables and the primary outcome (reaction time) allowing for the inclusion of adjustment for potential confounders (sleep duration, caffeine consumption). The primary variable of interest was a retrieval within the shift, included as a fixed effect. These models used a nested random effect (shift time within day and individual) to control with within individual correlation. Secondary outcomes were using the same modelling framework, only without adjustment. All analysis was completed using the R programming language.

Linear mixed effects regression ^26^ was performed on participant reaction time with a nested random effect structure of participant ID / day / shift (start vs. end), fixed effects included retrieval (dichotomous), Actigraph recorded sleep duration with adjustment for caffeine consumption during the shift. This model was reached following a step-down approach whereby all potential confounding variables were included in the model and were subsequently removed based on their collinearity and poor evidence of association with reaction time. A stepdown approach to regression was completed by initially including all listed adjusting variables in the model, which included duration and quality of sleep, age, gender, years’ experience, caffeine consumption and clinical role. Reaction time was the dependent variable.

The association between the Stanford sleepiness scale and reaction time was assessed using ordinal linear mixed regression. All analysis was completed using the R programming language. ^25-30^ Prior to analysis, all numeric values were shifted such that their mean was equal to 0 (i.e., variable centring), however, the scale of the values was left unchanged. **“**Invalid” reaction times, i.e., those that were < 100 or > 355 m**s**, false starts and lapses, respectively, were removed from the dataset.

## Results

The study participants were recruited between 2/1/2020 – 20/2/2021. Twenty-two participants were approached, with no exclusions. One participant withdrew without stating a reason, and two partially collected data owing to having trouble with sleep after wearing the wrist actigraph. Table 1 summarizes participant demographics and retrieval data. In total there were 194 lapses in reaction from 18 participants, and 1 false start.

**Table 1.** Participant demographics (mean (SD) unless otherwise stated)

|  |  |
| --- | --- |
| Age (years) | 34 (5.7) |
| Female: Male | 19:2 |
| Nurse: Doctor | 14:7 |
| Clinical experience (years) | 12 (4) |
| Total shifts assessed (all participants) | 79 |
| Total number of retrievals | 57 |
| Shifts with retrievals undertaken | 46 |
| Shifts with no retrieval undertaken | 33 |
| Retrieval time (hrs) * | 3 (2-4) |
| Mode of Transport (n) |  |
| Road | 53 |
| Fixed wing aircraft | 4 |
| Retrieval destination (n) |  |
| Metropolitan (Perth city area) | 51 |
| Remote (>70km) | 6 |
| Triage category | 2 (2) |

\*median (interquartile range)

### Effect of retrieval on RT

Mean RTs increased by 6.38 (95% CI: -2.17 to 14.92 ms) when a retrieval was undertaken, when keeping both sleep duration and caffeine consumption constant. The p value was 0.149, failing to demonstrate statistical significance, with the 95% confidence interval includes 0 change by a relatively small margin. Objectively (Actigraph) measured sleep (hrs) had no significant association with reaction time, with reaction time decreasing on average by 3.23 (95% CI: -8.08 to 1.62 ms) for each additional hour of sleep. Consuming caffeine during the shift increased mean reaction time by 16.26 (95% CI: 4.43 to 28.10 ms; p<0.009). The mean effect estimate of retrieval, objectively measured sleep, and caffeine consumption during the shift on RT are represented in Figure 1.

**Figure 1.**
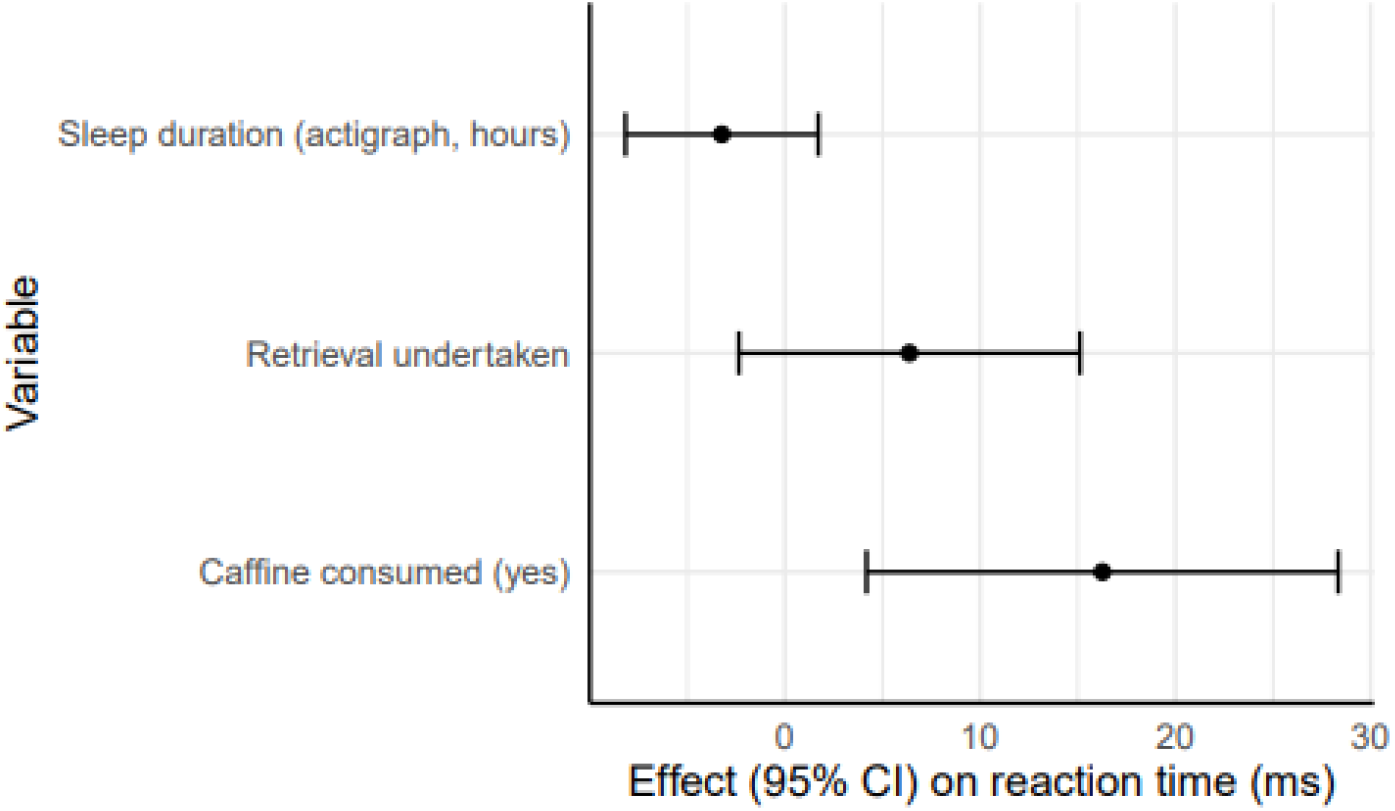
Effect estimates of dependant variable (95% confidence intervals: 95% CI).

### RT and the Stanford Sleepiness scale

When comparing RT to the Stanford Sleepiness Scale, as RT increased by 1 ms the odds of being in a more fatigued category increased by 0.57% (OR: 1.0057, 95% CI: 1.0036 to 1.0078). Confidence limits increase as the level of sleepiness increase suggesting that test precision is lost with increasing level of subjective sleepiness. The mean reaction times observed for each level in the Stanford sleepiness scale are seen in figure 2.

**Figure 2.**
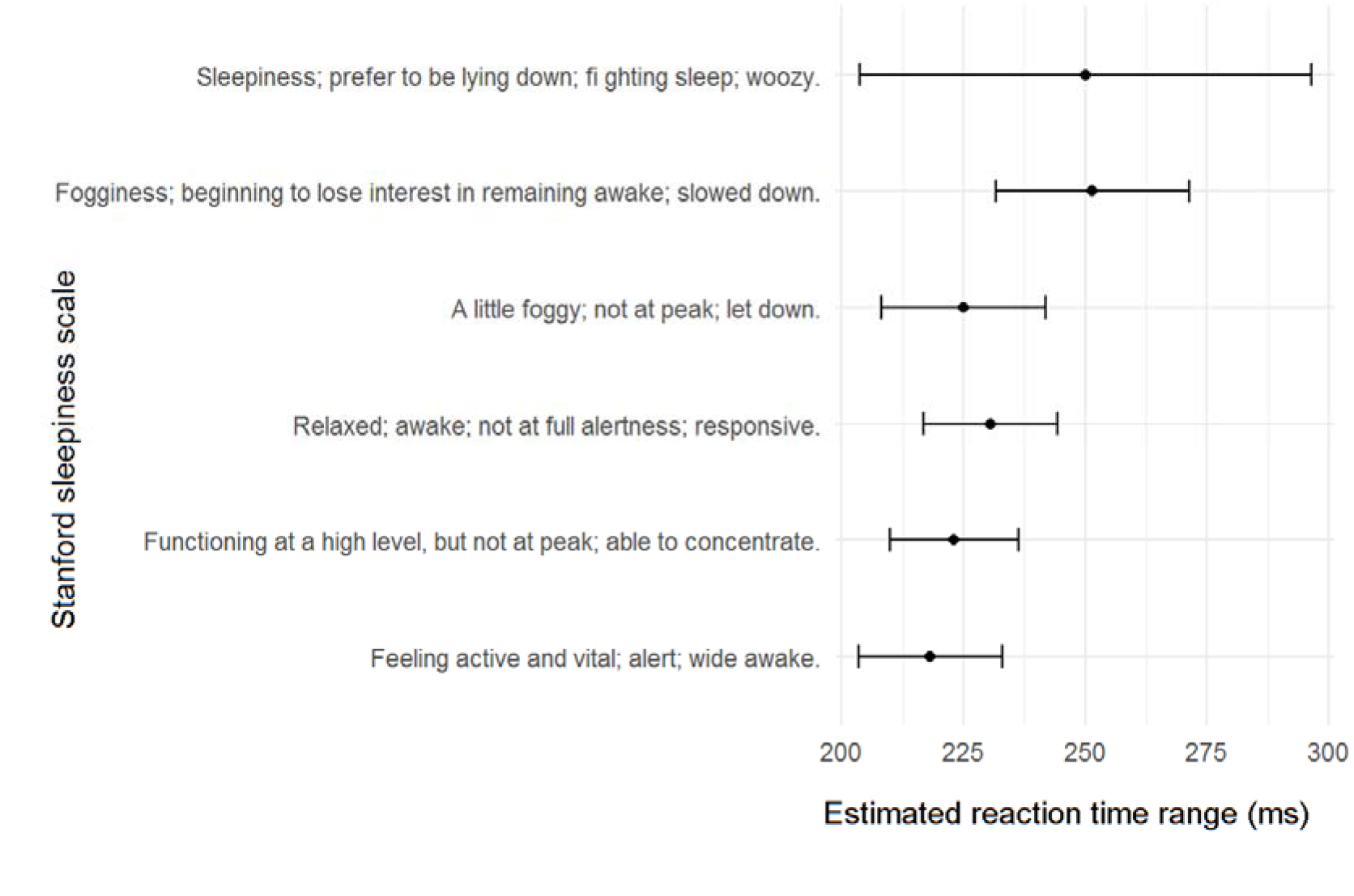
Mean reaction times observed for each level in the Stanford sleepiness scale. Estimates and 95% confidence intervals were determined by linear mixed effects modelling of reaction time vs sleepiness.

### Estimated hours of sleep correlates to objectively measured sleep

There was a positive correlation between subjective and objective sleep duration, for every extra hour of Actigraph sleep, subjective sleep increased by 0.18 hours (95% CI: 0.04 to 0.33 hours, approx. 11 min.). This means for 7hrs of self-reported sleep, actual number of hours slept is likely closer to 6hrs.

## Discussion

This novel study looked at the effect of the retrieval environment on psychometric vigilance as a measure of fatigue. To our knowledge, this is perhaps the first study measuring fatigue specifically in the neonatal retrieval environment. Our results showed that the 3-minute PVT via the *sleep-2-peak app* (Neuro Summum, Quebec, Canada) was easy to use in the field. Performing in a clinical retrieval prolonged reaction time but did not reach statistical significance. Sleep improved whereas caffeine consumption during a shift prolonged the RT. Subjective sleepiness was associated with a slower RT and participant’s estimated sleep duration correlated with objective recordings.

It appears counterintuitive that Caffeine prolonged the RT, but being an observational finding we cannot determine any causality. A recent meta-analysis reported caffeine improved psychometric vigilance in Emergency Medical Services Personnel, but also reduces sleep quality and quantity ^31^. This analysis was based on low-moderate quality of evidence. With its impact on sleep, there remains a paucity in evidence regarding how caffeine affects performance over time, with a need for studies to further investigate timing of caffeine consumption. To our knowledge, there are no reports on the use of effect of caffeine on reaction times in the retrieval environment. Given the potential benefits the effects of coffee consumption on RT and vigilance in a retrieval setting need further large studies.

Our results showed a positive correlation between subjective and objective sleep. However, participants tended to overestimate their sleep, a finding that is supported by other studies, with one study subjectively reporting sleep times averaging 6.8hrs and measured 6hrs^32^. In our study the average self-reported sleep was 7hrs (SD 0.75) and measured sleep 6.13hrs (SD 1.6)^32,33^

The significant association between subjective sleepiness and RTs is reported in the literature.^14,34-36^. Results of studies using self-reported tools should be interpreted cautiously ^37^. Our study shows poor reliability at higher levels of fatigue. Despite their limitations, self-reported surveys are an important tool to understand underlying cognitive processes in fatigue, since participants can let us know how their performance is being affected.^37^

In our study the 3-min PVT was a useful tool as an objective measure of fatigue in the neonatal retrieval environment. The authors of this study believe the impact of this study is to support the routine measurement of the 3-min PVT as part of a more rigorous approach to fatigue management in the retrieval environment. Whilst there are no clear thresholds for safety-critical performance for PVT, there is a growing body evidence aiming to identify these parameters and guide decision making regarding individual RTs and lapses ^8,38,39^. However, these studies as well as ours demonstrate that through regularly measuring PVT, subjective fatigue levels and sleep, crew members improve at self-assessment of their fatigue levels and understanding how to improve performance. This approach has the capacity to significantly enhance safety in aviation and transportation ^40^. In the aviation industry the Civil Aviation Safety Authority of Australia (CASA) stipulate operators have the choice of using prescriptive or flexible approaches ^41^. Hazard identification consists of predictive, proactive and reactive processes. Of the proactive measures both performance data (e.g., RTs) and fatigue surveys are recommended as well as sleep monitoring. There are also more sophisticated biomathematical models, such as the Sleep, Activity, Fatigue, & Task Effectiveness (SAFTE) model ^42^, which aims to simulate the physiological process of fatigue based on sleep and circadian rhythms. These are currently being adopted by the military and aviation industry. The medical industry has yet to establish such a rigorous approach to fatigue management.

## Conclusion

The 3-min PVT was found to be an easy and feasible method for objectively assessing fatigue in the neonatal retrieval setting. This study supports the literature in finding subjective sleepiness is not always a very reliable indicator of performance when fatigued. Larger studies are needed to further investigate long-range retrievals and the effects of caffeine. This study supports using the 3-min PVT routinely in combination with fatigue surveys and sleep monitoring as part of a more rigorous fatigue risk management system to improve patient and crew safety on neonatal transport.

## Data Availability

All data produced in the present study are available upon reasonable request to the authors

